# Metabolic Outcomes in Non-Alcoholic and Alcoholic Steatotic Liver Disease in among Korean and Adults: A Population-Based Multi-Cohort Study

**DOI:** 10.1101/2024.09.20.24313951

**Authors:** Yeongmin Kim, Taesic Lee, Chang-Myung Oh

## Abstract

**Background and aims:** This study investigated the prevalence and causal relationships of chronic metabolic diseases (diabetes, hypertension, and dyslipidemia) with steatotic liver disease (SLD), specifically metabolically associated alcoholic liver disease (MetALD).

**Methods:** We conducted a comprehensive analysis using cross-sectional data from the Korea National Health and Nutrition Examination Survey (KNHANES) from 2011 to 2021 and the National Health and Nutrition Examination Survey (NHANES) from 1999 to 2020. Longitudinal data from 2001 to 2014 from the Korean Genome and Epidemiology Study (KoGES) were used. Participants were categorized into the metabolic dysfunction-associated SLD (MASLD), MetALD, and ALD groups based on their hepatic steatosis index (HSI) and alcohol consumption. Logistic and Cox regression analyses were performed to assess the prevalence and incidence of chronic diseases.

**Results:** In both the KNHANES and NHANES cohorts, an increased HSI was significantly associated with a higher prevalence of chronic metabolic diseases. Longitudinal data from the KoGES cohort showed that MASLD and MetALD were significant predictors of chronic metabolic disease in both men and women. MetALD showed a higher hazard ratio for the development of chronic metabolic diseases than MASLD in Cox regression analysis.

**Conclusion:** This study highlighted the intertwined nature of SLD and metabolic health, with an emphasis on the role of MetALD. The significant association between MetALD and chronic metabolic diseases underscores the need for integrated management strategies that address both liver health and metabolic risk factors.

## 1. Introduction

Fatty liver disease (FLD) comprises a spectrum of liver disorders that is characterized by excessive accumulation of fat within hepatocytes. Non-alcoholic fatty liver disease (NAFLD), including non-alcoholic steatohepatitis, affects approximately 25% of adults worldwide^1^. NAFLD has emerged as a major health concern owing to its increasing prevalence and detrimental impact on global health^2^.

FLD is associated with various complications that significantly affect patient morbidity and mortality. Cardiovascular disease (CVD) is the leading cause of death in patients with FLD, reflecting the profound systemic effects of the disease^1, 2^. Additionally, patients with FLD are at an increased risk of developing extrahepatic malignancies, liver-related complications, chronic kidney disease, and type 2 diabetes mellitus^1^. These comorbidities illustrate the complex and multifaceted pathophysiology of FLD and underscore the need for a holistic and multidisciplinary approach to management and therapeutic interventions to mitigate the extensive burden of the associated comorbidities.

Recently, the scientific community revised the terminology from FLD to steatotic liver disease (SLD) and proposed a new classification of SLD to reflect a more nuanced understanding of its multifactorial etiology and complex pathophysiology. The newly proposed categories include metabolically associated SLD (MASLD), metabolically associated alcoholic liver disease (MetALD), and ALD^3^. MetALD includes patients with SLD due to mild alcohol consumption^3^. This refined classification system aims to better capture multiple metabolic and lifestyle factors that contribute to liver pathology, thereby facilitating more precise diagnosis and management strategies^3^.

Herein, we examined the associations and causal relationships between SLD and chronic metabolic disorders using community-based cohort data^4, 5^. We also analyzed the metabolic risks associated with MASLD, MetALD, and ALD in the context of chronic metabolic diseases using data from a comprehensive Korean cohort. By examining the prevalence and interplay of hypertension, diabetes, and dyslipidemia in these different liver disease categories, we aimed to elucidate their unique metabolic profiles, particularly MetALD. Our goal was to form clinical strategies for the management of patients with SLD and provide a basis for tailored healthcare approaches. Additionally, this study underscores the importance of accurate disease classification to address the broader burden of chronic metabolic diseases, ultimately contributing to improved patient care and outcomes.

## 2. Methods

### 2.1 Study Population

We conducted a comprehensive analysis using cross-sectional data from the 2011– 2021 Korea National Health and Nutrition Examination Surveys (KNHANES) and the 1999– 2020 National Health and Nutrition Examination Survey (NHANES). The KNHANES datasets are publicly available on the KNHANES website (http://knhanes.cdc.go.kr)^6^. NHANES records are publicly available on the CDC website (www.cdc.gov/nchs/nhanes/index.htm)^7^. All participants consented to the use of their information for research purposes. Both the KNHANES and NHANES datasets contain extensive information on health, nutrition, medical history, physical measurements, and laboratory results (Supplementary Figures 1 and 2). Additionally, we examined the longitudinal data from the Korean Genome and Epidemiology Study (KoGES) from 2001 to 2014 that included detailed records of health, diet, medical history, physical measurements, and laboratory results (Supplementary Figure 3)^4^. These datasets served as the basis for our investigation of the association between liver diseases, specifically variations related to alcohol consumption and chronic diseases such as diabetes, hypertension, and dyslipidemia.

The KNHANES and NHANES datasets were preprocessed and merged into a single file, regardless of the survey year, using R software version 4.1.2 (Figure 1A)^8^. Participants with missing values for key baseline characteristics, including systolic blood pressure (SBP), waist circumference (WC), height, weight, body mass index (BMI), triglycerides (TG), total cholesterol (TC), glucose, and hemoglobin A1c (HbA1c), were excluded from further analysis. Participants with missing values for variables used in the subsequent analyses, including aspartate aminotransferase (AST), alanine aminotransferase (ALT), alcohol consumption, and smoking status, were also excluded. Participants with missing data on diabetes, hypertension, dyslipidemia, and use of dyslipidemia medications were also excluded. The KoGES dataset underwent similar pre-processing, resulting in a single consolidated file. Participants with missing values for the same baseline characteristics were excluded from the analysis. Similarly, those with missing data on AST and ALT levels, alcohol consumption, and smoking status were excluded from further analyses. To accurately determine the presence of diabetes, hypertension, and dyslipidemia as well as the use of dyslipidemia medications, participants with missing data for these variables were excluded. To examine the causality of chronic diseases resulting from fatty liver disease, we developed separate models for diabetes, hypertension, and dyslipidemia. Only participants with at least seven follow-up records (over 12 years) were included in the causality analysis to ensure robust longitudinal data for reliable inferences.

**Figure 1.**
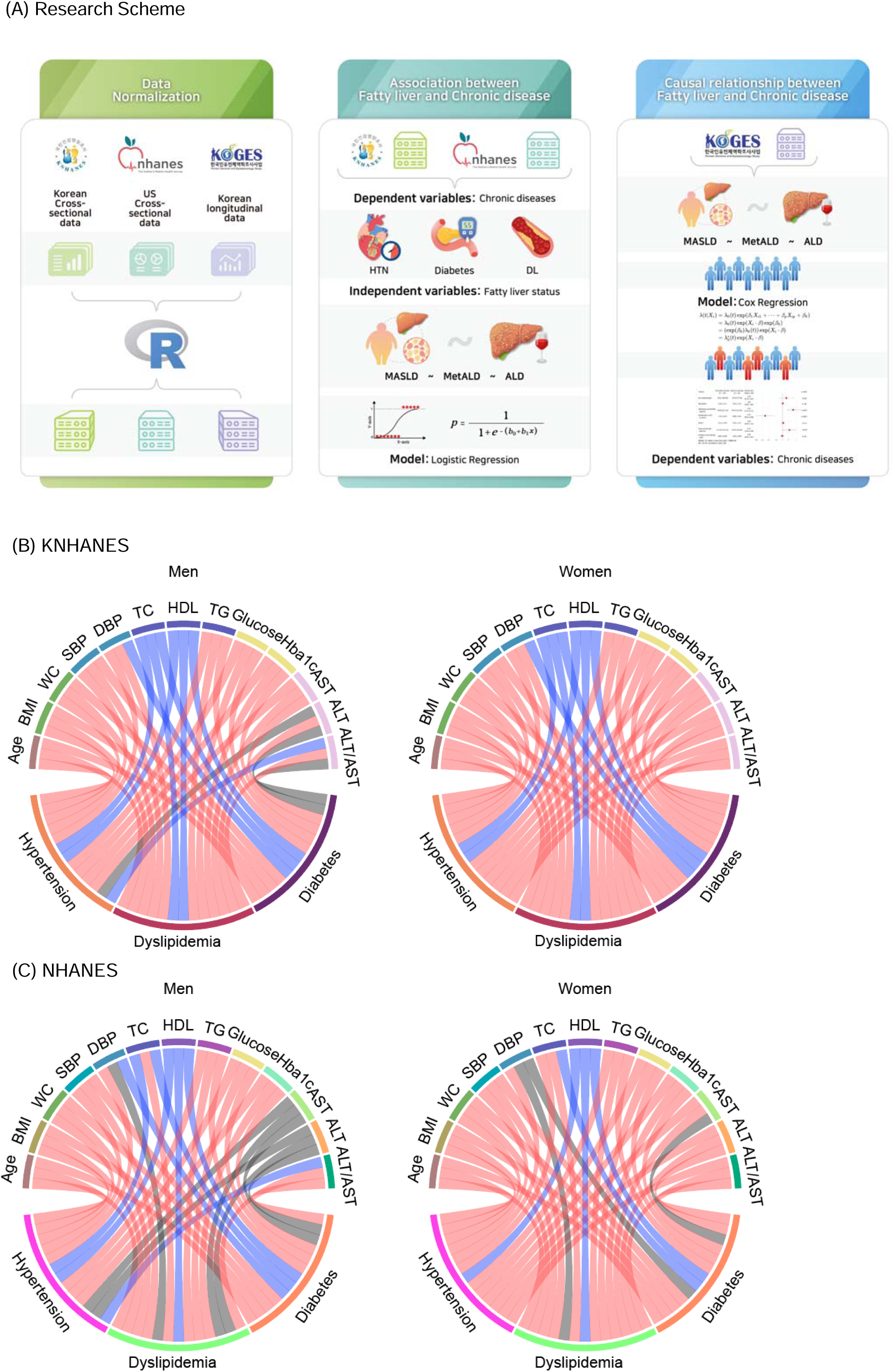
Associations between chronic metabolic disease and metabolic parameters. (A) Research scheme for data normalization, association, and causality analysis for the three cohorts. (B) Circos plot of associations between chronic metabolic diseases and metabolic parameters in the KNHANES cohort. (C) Circos plot of associations between chronic metabolic diseases and metabolic parameters in the NHANES cohort.

### 2.2 Definition of Chronic Diseases and MASLD

The presence of chronic diseases, including diabetes and hypertension, and the classification of healthy participants were determined based on whether a physician had diagnosed the disease at least once. Dyslipidemia was identified by either a physician’s diagnosis or the use of dyslipidemia medications. Fatty liver status was assessed using the hepatic steatosis index (HSI), which was calculated using the following formula: HIS = 8 × ALT/AST + BMI + 2 (if diabetic) + 2 (if female)^9^. A HSI score of ≥36 was used to diagnose fatty liver disease, as established in previous research^9–11^.

For participants with SLD, daily alcohol consumption was used to further categorize the participants. Men who consumed <30 g and women who consumed <20 g of alcohol per day were classified as having MASLD, whereas men who consumed between 30 g and 60 g and women who consumed between 20 g and 50 g of alcohol per day were classified as having MetALD. Finally, men who consumed >60 g and women who consumed >50 g of alcohol per day were classified as having ALD^3^.

### 2.3 Collection of Data and Covariates

Demographics of participants with and without liver disease across were obtained from KNHANES, NHANES, and KoGES. Participants were stratified based on the presence or absence of liver disease. Demographic data are presented as mean ± standard deviation for continuous variables and as frequency and percentage (%) for categorical variables. To select the covariates, we conducted a comprehensive review of eight peer-reviewed studies^11–18^ that used logistic regression or Cox regression models with covariates. These studies were selected based on their relevance to the analysis of covariates in the context of fatty liver disease and chronic diseases. The number of covariates used in these studies were 24 variables (Supplementary Table 2). The table also shows the covariates included in the multivariate analyses of each study.

To investigate the association between SLD and chronic diseases, we included variables related to liver damage, specifically AST and ALT levels, ALT/AST ratio, HSI, and alcohol consumption. Each of these variables was treated as continuous, and values were categorized into eight groups to examine the proportion of chronic diseases in each group.

For alcohol consumption, different grouping criteria were applied based on sex in accordance with MASLD guidelines and as follows: men were grouped into 0–30, 30–60, and >60 g per day, while women were grouped into 0–20, 20–50, and >50 g per day.

### 2.4 Statistical analysis

To assess the statistical significance of the differences in the characteristics between groups with and without chronic diseases, one-way analysis of variance was employed, and p-values were reported. Logistic and Cox regression analyses were performed to determine the prevalence and incidence of chronic diseases related to SLD. Multivariate logistic regression was used to calculate the odds ratios (ORs) for chronic disease prevalence, whereas Cox regression was used to calculate the hazard ratios (HRs) for chronic disease incidence after adjusting for selected covariates. All statistical analyses were conducted using R software version 4.1.2, and significance was set at p < 0.05.

### 2.5 Ethical considerations

The Institutional Review Board of Gwangju Institute of Science and Technology (South Korea) approved the study protocol (IRB No. 20221201-BR-69-02-02). All research procedures were conducted in accordance with the relevant guidelines and regulations. All participants volunteered and provided written informed consent before enrollment, and their records were anonymized before being accessed by the authors.

All authors had access to the study data and reviewed and approved the final manuscript

## 3. Results

### 3.1 Baseline characteristics of the participants

The general characteristics of the participants in the KNHANES, NHANES, and KoGES cohorts according to fatty liver status are summarized in Tables 1, 2, and Supplementary Table 1. The KNHANES cohort included 22,597 healthy men and 7,017 men with fatty liver. Among men with fatty liver, most were categorized under MASLD (n = 3,494), followed by ALD (n = 2,980) and MetALD (n = 543) (**Table 1**). Meanwhile, there were 29,493 healthy women and 7,042 women with fatty liver, primarily with MASLD (n = 5,733) (**Table 1**). Healthy Korean men were generally older, whereas those with fatty liver were younger; no substantial age differences were observed among the MASLD, MetALD, and ALD groups. For women, no differences were observed among healthy subjects and those with MetALD and ADL. Women with MASLD were generally older than men with MASLD.

**Table 1.** General characteristics of the participants in the KNHANES cohort according to fatty liver status.

|  | Healthy | Fatty Liver Disease |  |  |
| --- | --- | --- | --- | --- |
|  |  | MASLD | MetALD | ALD |
| Male, N | 22597 | 3494 | 543 | 2980 |
| Age, years | 47.18 ± 20.824 | 43.74 ± 18.147 | 43.42 ± 14.546 | 44.24 ± 13.415 |
| BMI | 22.8 ± 2.731 | 27.93 ± 3.127 | 27.86 ± 3.331 | 28.21 ± 2.871 |
| Waist circumference | 81.6 ± 9.093 | 94.4 ± 8.357 | 94.15 ± 8.85 | 95.42 ± 7.677 |
| Systolic blood pressure, mm Hg | 119.04 ± 15.394 | 121.49 ± 13.183 | 121.91 ± 13.157 | 124.18 ± 13.477 |
| Diastolic blood pressure, mm Hg | 74.84 ± 10.724 | 78.68 ± 10.352 | 80.34 ± 10.424 | 82.68 ± 10.569 |
| Total cholesterol, mg/dL | 182.02 ± 36.447 | 189.38 ± 39.178 | 195.06 ± 40.681 | 197.72 ± 39.084 |
| HDL cholesterol, mg/dL | 49.19 ± 11.534 | 42.07 ± 8.56 | 42.76 ± 8.174 | 45.11 ± 9.862 |
| Triglyceride, mg/dL | 133.35 ± 114.166 | 171.65 ± 120.11 | 191.9 ± 142.22 | 220.84 ± 165.85 |
| Fasting plasma glucose, mg/dL | 99.96 ± 21.363 | 108.16 ± 30.403 | 105.82 ± 24.338 | 110.41 ± 30.367 |
| HbA1C, % | 5.69 ± 0.746 | 6.03 ± 1.078 | 5.9 ± 0.873 | 5.97 ± 1 |
| AST, IU/L | 23.9 ± 15.745 | 28.54 ± 16.338 | 28.84 ± 20.829 | 29.55 ± 16.02 |
| ALT, IU/L | 20.43 ± 12.199 | 45.18 ± 34.874 | 45.3 ± 38.737 | 43.89 ± 31.044 |
| ALT/AST | 0.86 ± 0.251 | 1.53 ± 0.41 | 1.54 ± 0.413 | 1.45 ± 0.389 |
| HIS | 29.87 ± 3.778 | 40.49 ± 4.149 | 40.4 ± 4.372 | 40.09 ± 3.771 |
| Alcohol Intake, g/day | 100.37 ± 159.501 | 6.04 ± 8.416 | 40.87 ± 2.787 | 248.69 ± 182.875 |
| Female, N | 29493 | 5733 | 650 | 659 |
| Age, years | 46.62 ± 19.29 | 55.67 ± 15.976 | 47.87 ± 14.192 | 44.81 ± 13.945 |
| BMI | 22.06 ± 2.646 | 28.21 ± 3.214 | 28.6 ± 3.201 | 28.85 ± 3.472 |
| Waist circumference | 75.44 ± 8.442 | 90.89 ± 8.332 | 90.95 ± 8.3 | 91.94 ± 8.725 |
| Systolic blood pressure, mm Hg | 114.32 ± 17.064 | 124.19 ± 16.912 | 120.69 ± 15.933 | 121.43 ± 16.226 |
| Diastolic blood pressure, mm Hg | 71.96 ± 9.507 | 76.1 ± 9.663 | 77.71 ± 9.699 | 78.47 ± 9.929 |
| Total cholesterol, mg/dL | 188.75 ± 36.134 | 193.94 ± 40.556 | 197.3 ± 42.156 | 199.38 ± 40.422 |
| HDL cholesterol, mg/dL | 55.4 ± 12.53 | 48.48 ± 10.732 | 50.21 ± 11.096 | 53.15 ± 12.519 |
| Triglyceride, mg/dL | 103.57 ± 69.52 | 149.06 ± 91.456 | 148.85 ± 100.813 | 165.37 ± 137.577 |
| Fasting plasma glucose, mg/dL | 94.7 ± 16.392 | 111.18 ± 32.65 | 107.16 ± 26.98 | 108.42 ± 28.447 |
| HbA1C, % | 5.59 ± 0.59 | 6.24 ± 1.109 | 5.99 ± 0.962 | 5.87 ± 0.944 |
| AST, IU/L | 20.39 ± 9.815 | 25.25 ± 13.301 | 24.83 ± 15.091 | 26.4 ± 17.383 |
| ALT, IU/L | 15.16 ± 8.607 | 29.45 ± 21.934 | 30.42 ± 29.446 | 31.4 ± 29.653 |
| ALT/AST | 0.74 ± 0.196 | 1.13 ± 0.32 | 1.15 ± 0.335 | 1.15 ± 0.5 |
| HIS | 30.05 ± 3.313 | 39.7 ± 3.503 | 40.09 ± 3.55 | 40.21 ± 4.561 |
| Alcohol Intake, g/day | 25.03 ± 72.955 | 2.12 ± 3.498 | 33.04 ± 8.483 | 202 ± 166.539 |

Data are presented as mean ± standard deviation.
HDL, high-density lipoprotein; AST, aspartate transaminase; ALT, alanine transaminase; HIS, hematocrit

**Table 2.**
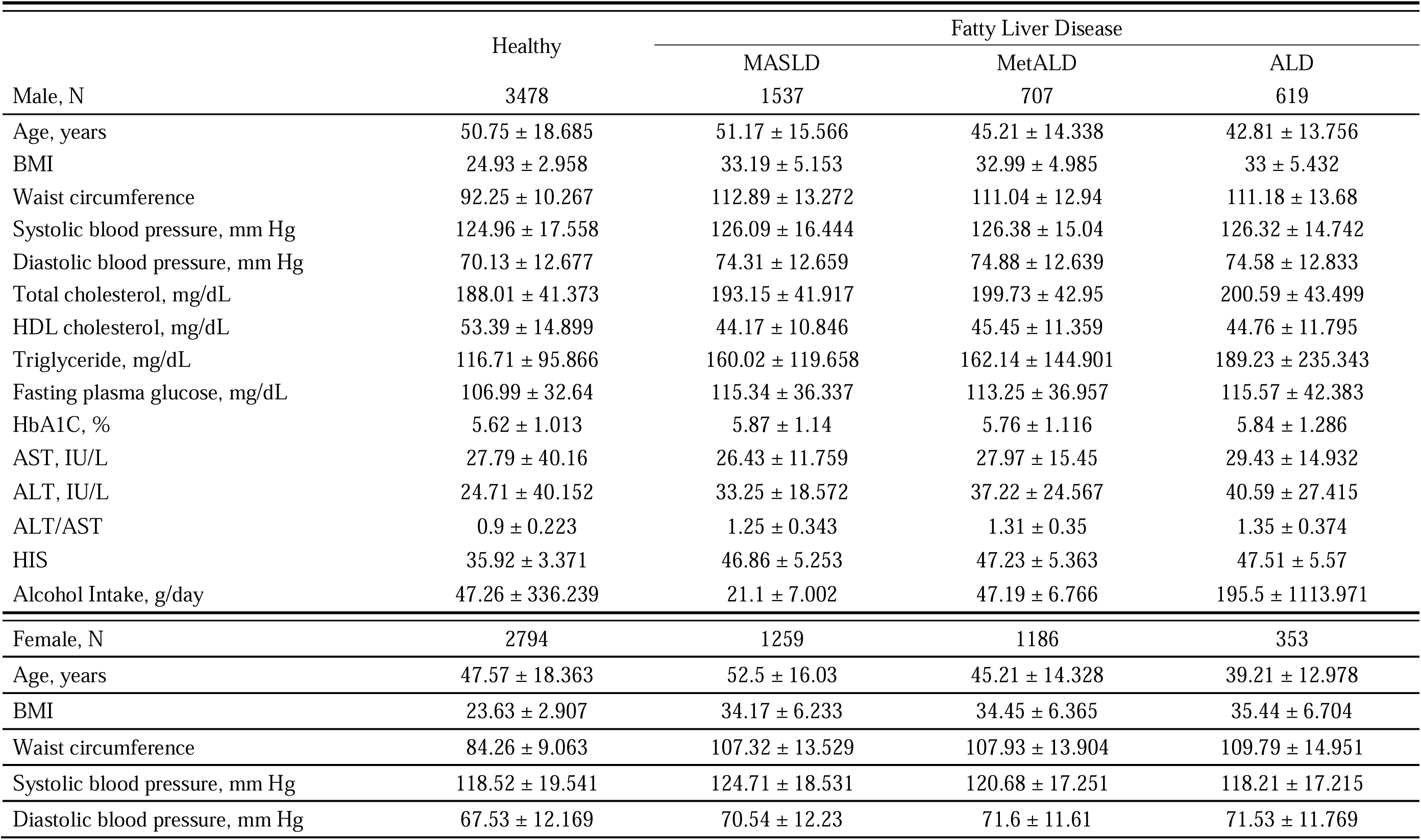
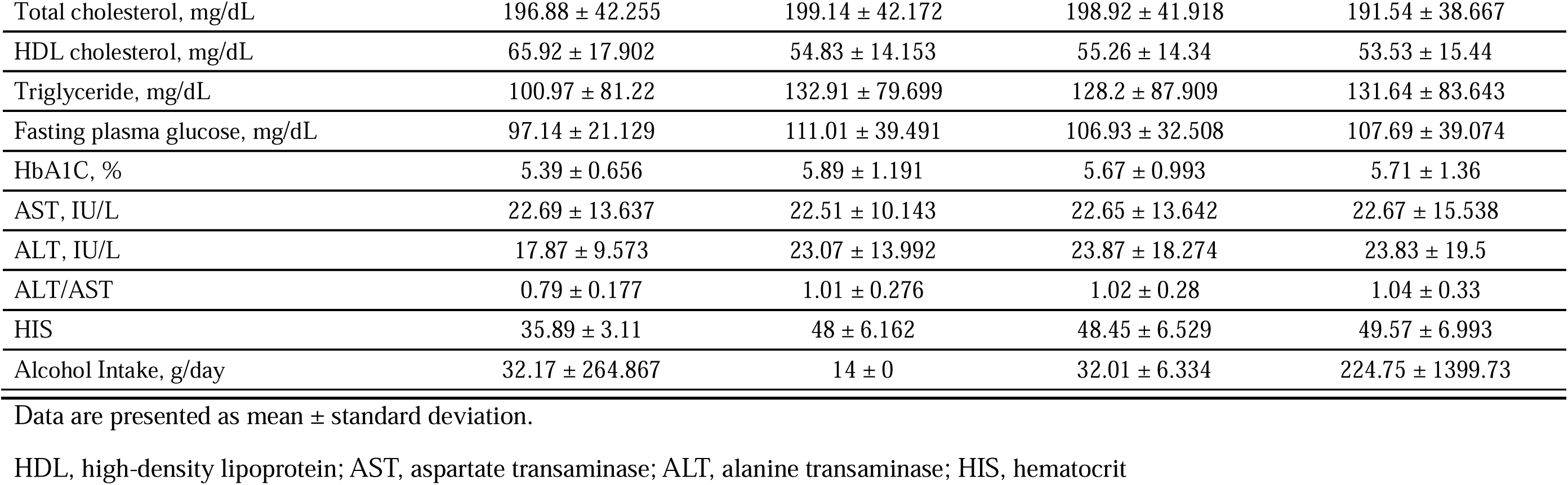
General characteristics of the participants in the NHANES cohort according to fatty liver status.

There are no established HSI thresholds for determining the presence of fatty liver in ethnic groups outside Korea. Therefore, we adopted an HSI threshold of 41 for subsequent analyses for two reasons. First, we approximated the prevalence in the Korean population (Supplementary Figure 4). Second, we adjusted for the BMI difference, which was approximately 5, between the KNHANES and NHANES cohorts. The NHANES cohort included 3,478 healthy men and 2,863 men with fatty liver. Among men with fatty liver, most were categorized under MASLD (n = 1,537), followed by MetALD (n = 707) and ALD (n = 619) (**Table 2**). Meanwhile, there were 2,794 healthy women and 2,798 women with fatty liver, primarily with MASLD (n = 1,259) (**Table 2**). Men with MetALD and ALD were generally younger, similar to those in the KNHANES cohort. Women with MASLD and ALD were generally older and younger, respectively, similar to those in the KNHANES cohort.

The KoGES cohort included 3,235 healthy men, 744 men with MASLD, 98 with MetALD, and 72 with ALD. Meanwhile, there were 3,273 healthy women, 1,291 women with MASLD, 26 with MetALD, and 5 with ALD. In the KNHANES, NHANES, and KoGES cohorts, men with MetALD and ALD were generally younger, whereas those with MASLD varied. Women with MASLD were generally older and had younger ALD patterns.

Across all cohorts, participants with fatty liver had significantly higher BMI, WC, and TC levels than healthy subjects regardless of sex. Markers of liver injury, such as AST and ALT levels, were elevated in patients in the MASLD, MetALD, and ALD groups compared to healthy participants. The ALT/AST ratio was also consistently increased in patients in the MASLD, MetALD, and ALD groups compared with healthy participants. Blood parameters associated with hypertension, such as SBP and diastolic BP (DBP), were raised in subjects with fatty liver compared with healthy subjects in all cohorts and regardless of sex. Dyslipidemia parameters, including TC levels, were higher in participants with fatty liver than in healthy individuals regardless of sex. Conversely, high-density lipoprotein (HDL) levels were lower in participants with fatty liver than in healthy participants in all cohorts and regardless of sex sexes. Additionally, diabetes parameters, including fasting glucose and HbA1c levels, were elevated in participants with fatty liver compared to healthy participants in all cohorts and regardless of sex.

### 3.2 Associations between chronic metabolic disease and disease parameters

The prevalence of chronic diseases, such as diabetes, hypertension, and dyslipidemia, increase annually^19–21^. Tracking annual prevalence trends using preprocessed cross-sectional data from the NHANES and KNHANES databases revealed an increasing prevalence of diabetes, hypertension, and dyslipidemia over time (Supplementary Figure 5).

Sex-specific association patterns between chronic metabolic diseases and clinical parameters were compared between the two cross-sectional cohorts. In the KNHANES cohort (Figure 1B), hypertension in both sexes showed positive correlations with age, BMI, WC, SBP, DBP, and TG, fasting glucose, HbA1c, and AST levels and negative linear trends with TC and HDL levels. When comparing the sex-specific hypertension-related signatures in the KNHANES and NHANES cohorts, the following variables exhibited correlations independent of sex and ethnicity: age, BMI, WC, SBP, DBP, TG, fasting glucose, and HbA1c levels. In Korea, the pattern of association between liver profile and hypertension risk differed between men and women but was consistently positive in men and women in the United States. TC and HDL levels were negatively related to the prevalence of hypertension in most cases, except for women in the United States.

Dyslipidemia-related signatures largely coincide with those linked to hypertension risk, including TG, fasting glucose, and ALT levels, as well as the ALT/AST ratio, with the exception of DBP and TC. The ALT/AST ratio, which is a predictor of NAFLD and IR^9, 22^ exhibited a positive relationship with the ratio of dyslipidemia regardless of sex and ethnicity. Furthermore, TC levels showed a positive association with the risk of dyslipidemia in the US dataset and a negative association in the Korean cohort.

Diabetes-related signatures mostly converged with those associated with the risk of diabetes, except for DBP and the ALT/AST ratio, which exhibited a positive relationship with the hypertension ratio depending on sex.

### 3.3 Associations between chronic metabolic disease and hepatic steatosis in the KNHANES cohort

HSI of patients in the KNHANES cohort were transformed into octile percentiles to identify linear trends in hepatic steatosis status with the prevalence of chronic diseases. As HSI increased, all diseases exhibited monotonic escalation patterns (Figures 2A, 2B).

**Figure 2.**
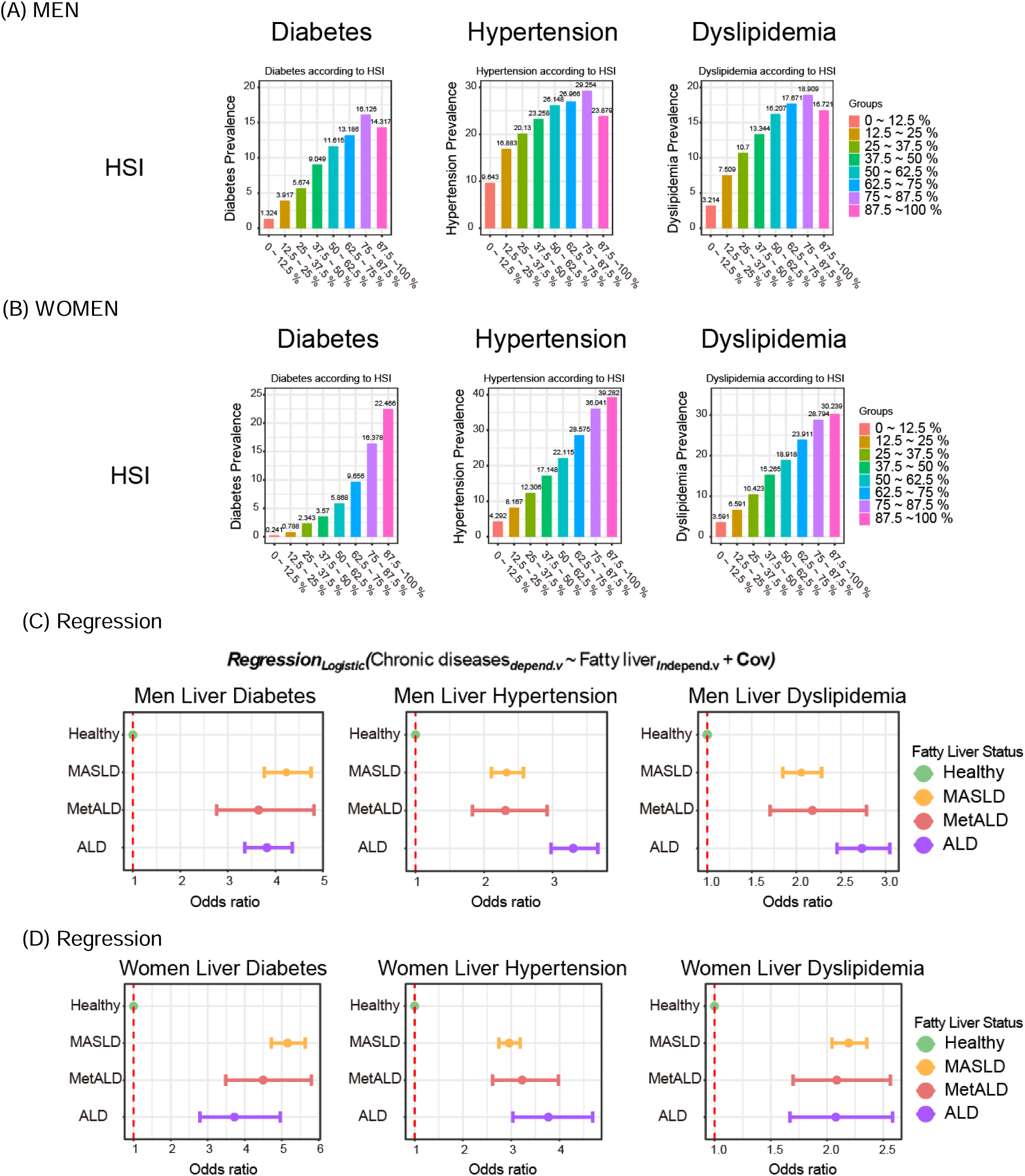
Disease prevalence according to hepatic steatosis in the KNHANES cohort. (A, B) Chronic metabolic disease prevalence according to the hepatic steatosis index (HSI) in men (A) and women (B). (C, D) Hazard ratio of chronic metabolic disease according to fatty liver status in multiple logistic regression analysis in men (C) and women (D). MASLD, metabolic dysfunction-associated steatotic liver disease; MetALD, metabolically associated alcoholic liver disease; ALD, alcoholic liver disease.

After an extensive review of existing literature, age and smoking status were identified as important confounding factors that could significantly affect the relationship between fatty liver status and chronic diseases (Supplementary Table 2). The association between fatty liver status and chronic diseases was further assessed using a multivariable logistic regression model that adjusted for confounders such as age and smoking status. Among men in the KNHANES cohort, the ORs for MASLD, MetALD, and ALD were 4.232 (95% CI, 3.767–4.755), 3.647 (95% CI, 2.764–4.813), and 3.824 (95% CI, 3.356–4.357) for diabetes (Figures 2C, 2D), 2.330 (95% CI, 2.108–2.576), 2.312 (95% CI, 1.829–2.921), and 3.301 (95% CI, 2.977–3.660) for hypertension, and 2.055 (95% CI, 1.848–2.285), 2.180 (95% CI, 1.706–2.787), and 2.735 (95% CI, 2.456–3.046) for dyslipidemia, respectively. Among women in the KNHANES cohort, the ORs for diabetes (MASDL > MetALD > ALD), hypertension (ALD > MetALD > MASLD), and dyslipidemia (MASLD > MeALD > ALD) were similar to those in men.

### 3.4 Associations between chronic metabolic disease and hepatic steatosis in the NHANES cohort

HSI in the NHANES data were transformed into octile percentiles to identify linear trends in hepatic steatosis status with the prevalence of chronic diseases. The results showed that as HSI increased, all diseases exhibited monotonic escalation patterns (Figures 3A, 3B).

**Figure 3.**
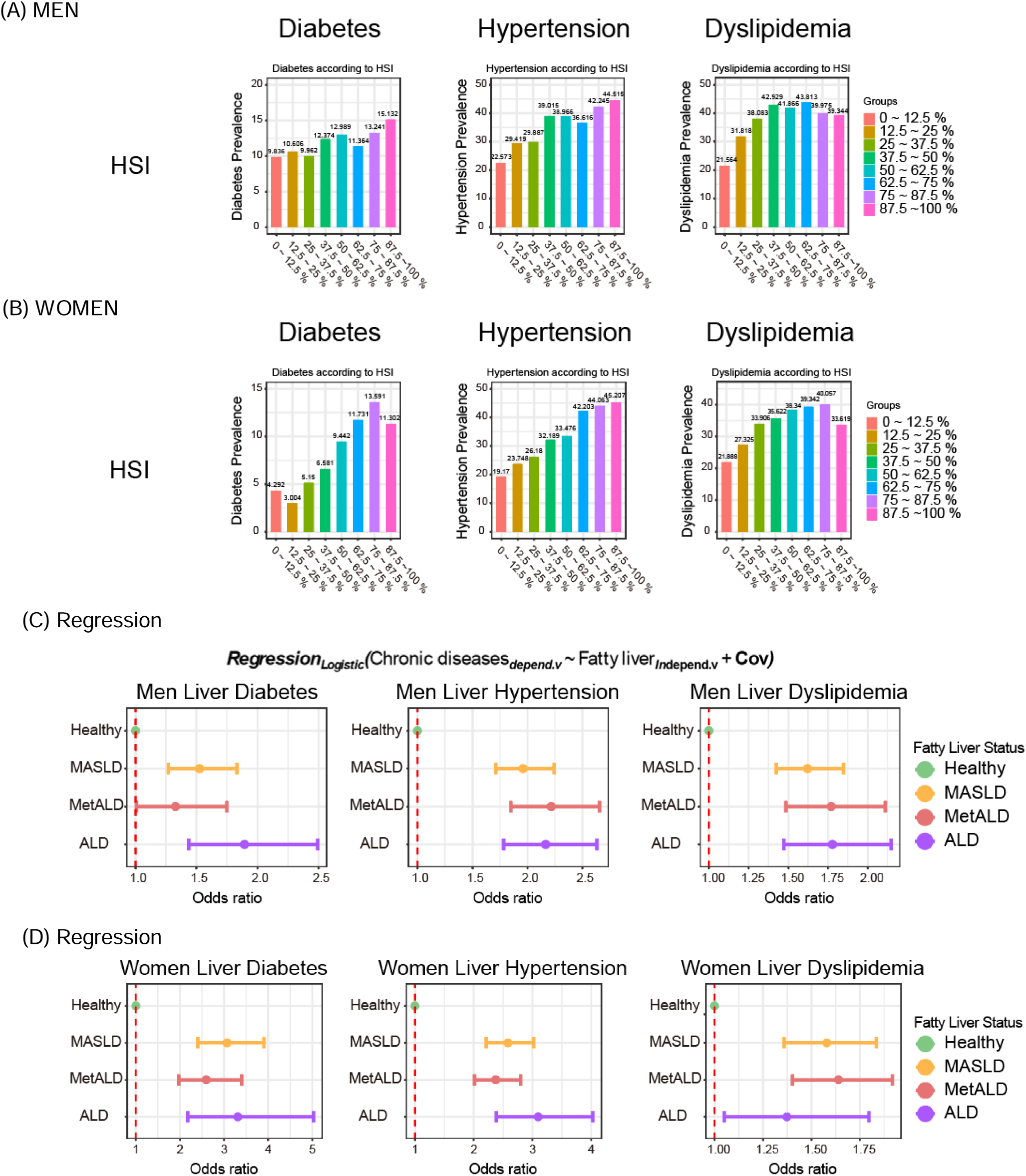
Disease prevalence according to hepatic steatosis in the NHANES cohort. (A, B) Chronic metabolic disease prevalence according to the hepatic steatosis index (HSI) in men (A) and women (B). (C, D) Hazard ratio of chronic metabolic disease according to fatty liver status in multiple logistic regression analysis in men (C) and women (D). MASLD, metabolic dysfunction-associated steatotic liver disease; MetALD, metabolically associated alcoholic liver disease; ALD, alcoholic liver disease.

The association between fatty liver and chronic diseases was assessed using a multivariate logistic regression model. Among men in the NHANES cohort, the ORs for MASLD, MetALD, and ALD were 1.524 (95% CI: 1.269–1.830), 1.328 (95% CI: 1.009–1.748), and 1.893 (95% CI: 1.438–2.492) for diabetes (Figures 3C, 3D), 1.960 (95% CI, 1.713–2.242), 2.215 (95% CI, 1.848–2.656), and 2.166 (95% CI, 1.783–2.631) for hypertension, and 1.621 (95% CI, 1.423–1.846), 1.770 (95% CI, 1.484–2.111), and 1.778 (95% CI, 1.472–2.147) for dyslipidemia, respectively. Among women in the NHANES cohort, the ORs for diabetes, hypertension (ALD > MASDL > MetALD), and dyslipidemia (MetALD > MASLD > ALD) were similar to those in men.

We also analyzed the association between alcohol consumption and the prevalence of chronic metabolic diseases and found no significant increase in the KNHANES or NHANES cohorts. However, in the KNHANES cohort, an inverse relationship was observed among women, wherein higher alcohol consumption was associated with a lower prevalence of chronic metabolic diseases (Supplementary Figure 6).

### 3.5 Risks of Chronic Metabolic Diseases in SLD

To examine the causal relationship between chronic diseases and fatty liver status, we used longitudinal data from the KoGES cohort and performed Cox regression analysis. Figure 4 shows the causal effects of the fatty liver status on the development of chronic metabolic diseases. To predict the development of chronic metabolic diseases in patients with fatty liver disease, we used data from 2 years.

**Figure 4.**
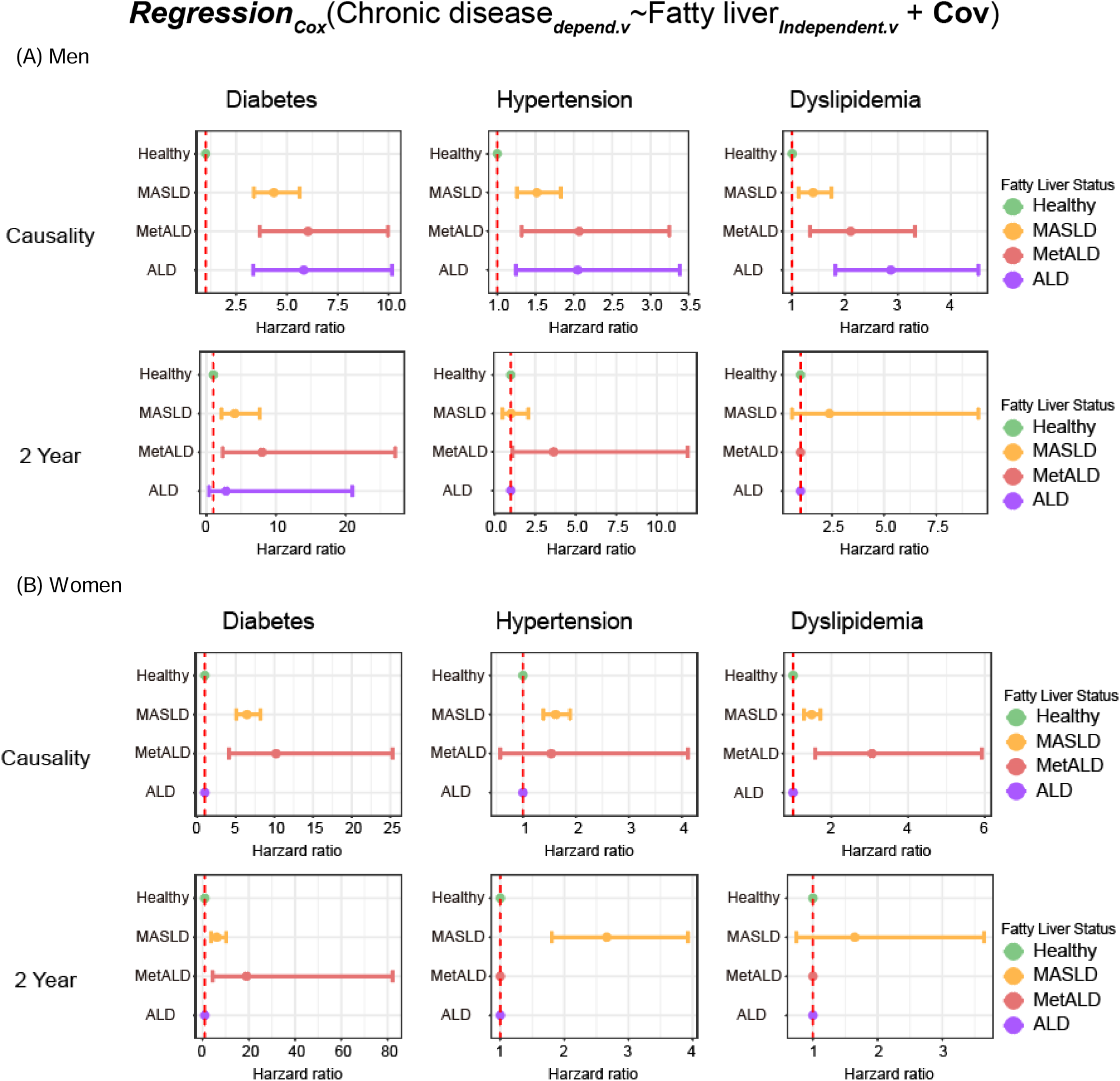
Causal relationship between chronic metabolic disease and fatty liver status in the KoGES cohort. (A) Hazard ratio of chronic metabolic disease according to fatty liver status in Cox regression analysis in men. (B) Hazard ratio of chronic metabolic disease according to fatty liver status in Cox regression analysis in women. Causality: Cox regression analysis was performed to assess the relationship between baseline fatty liver status and the incidence of chronic metabolic diseases over the 2-year follow-up period.

Based on the 2-year data, we calculated the risk of early development of chronic metabolic diseases. Both MASLD and MetALD were significant predictors of diabetes in men (HR, 4.076; 95% CI, 2.172–7.647 and HR, 8.012; 95% CI, 2.365–27.139, respectively) and women (HR, 6.450; 95% CI, 5.072–8.202 and HR, 10.218; 95% CI, 4.130–25.285, respectively). For hypertension, MetALD was a significant predictor in men (HR, 3.637; 95% CI, 1.111–11.900), whereas MASLD was a significant predictor in women (HR, 2.662; 95% CI, 1.803–3.930). For dyslipidemia, the 2-year data showed no significant predictive value based on fatty liver status in either sex (Figures 4A, 4B).

Regarding overall causality, among men (Figure 4A), the HRs for MASLD, MetALD, and ALD were 4.354 (95% CI, 3.371–5.624), 6.037 (95% CI, 3.654–9.975), and 5.831 (95% CI, 3.343–10.171) for diabetes, 1.517 (95% CI, 1.257–1.830), 2.069 (95% CI, 1.319–3.245), and 2.050 (95% CI, 1.242–3.383) for hypertension, and 1.398 (95% CI, 1.120–1.743), 2.109 (95% CI, 1.338–3.325), and 2.867 (95% CI, 1.818–4.522) for dyslipidemia, respectively.

Meanwhile, the number of women with ALD was insufficient to provide reliable HR estimates; however, their HRs for diabetes and dyslipidemia were similar to those in men. Additionally, their HRs for hypertension were significant only for MASLD.

## 4. Discussion

In this study, we comprehensively examined the prevalence and causality of chronic metabolic diseases, such as diabetes, hypertension, and dyslipidemia, in relation to fatty liver status. We used cross-sectional data from KNHANES and NHANES and longitudinal data from KoGES. Among men, the association between chronic metabolic diseases and liver injury markers was inconsistently significant. However, in women, positive correlations were observed between these conditions and AST and ALT levels, suggesting a stronger association between liver injury and chronic conditions (Figures 1B, 1C). The prevalence of chronic diseases also significantly increased with HSI in both men and women in the KNHANES and NHANES cohorts (Figures 2A, 2B, 3A, and 3B), highlighting the critical role of liver health in the management of metabolic diseases. Logistic regression analysis confirmed these associations, with higher HSI correlating with an increased risk of chronic metabolic disease (Figures 2C, 2D, 3C, and 3D). MetALD emerged as a significant predictor, underscoring the need for further attention to this newly defined category.

A comparison of the two cohorts revealed some notable differences. In the KNHANES cohort, the prevalence of chronic metabolic diseases did not increase with alcohol consumption. Interestingly, higher alcohol consumption was associated with a lower prevalence of these diseases among women, suggesting a possible protective effect of moderate alcohol consumption. However, this was not observed in the NHANES cohort (Supplementary Figure 6), suggesting possible differences in the population characteristics or lifestyle factors between Korean and American adults.

Using longitudinal data from the KoGES cohort, we performed Cox regression analyses to explore causal relationships (Figure 4). MetALD was a significant predictor of diabetes in both men and women, with the HRs indicating a strong association. MetALD is a significant predictor of hypertension in men, highlighting its role in the progression of cardiovascular diseases. Predictions for dyslipidemia varied, with MetALD showing significant associations in both men and women, although the magnitudes were different. The unique metabolic profile of MetALD underscores the importance of its recent categorization. This group, which is characterized by specific alcohol consumption patterns coupled with metabolic risk factors, highlights the complex interplay between lifestyle factors and liver health. Our findings suggest that MetALD is not just an intermediate category but a distinct entity with specific clinical implications.

This study had several limitations. First, the cross-sectional nature of the KNHANES and NHANES data limits their ability to establish causality. Although longitudinal data from the KoGES cohort provided insights into causal relationships, the length of follow-up may not fully capture the long-term progression of liver disease and chronic metabolic conditions. Second, the reliance on self-reported data on alcohol consumption and other lifestyle factors may introduce recall bias, potentially affecting the accuracy of categorization into MASLD, MetALD, and ALD.

In conclusion, this study underscores the intertwined nature of liver and metabolic health, with a particular emphasis on the role of MetALD. The significant association between MetALD and chronic metabolic diseases highlights the need for integrated management strategies that address both liver health and metabolic risk factors. Future research should continue to explore the specific mechanisms underlying MetALD to further refine diagnostic and therapeutic approaches.

## Supporting information

Supplementary Figure.docx

## Data Availability

KoGES
NHANES
KNHAENES

## Declaration

### CRediT Authorship Contributions

Chang-Myung Oh (Writing – original draft: Supporting; Writing – review and editing: lead). Taesic Lee (Data curation: Lead; Writing – review and editing: support) Yeongmin Kim (Data curation: Supporting; Writing: original draft: Lead; Writing: review and editing: Supporting)

### Ethics approval and consent to participate

This study was approved by the Institutional Review Board (IRB) of Gwangju Institute of Science and Technology (20221201-BR-69-02-02).

