## Supplementary Figure.docx for "Metabolic Outcomes in Non-Alcoholic and Alcoholic Steatotic Liver Disease in among Korean and Adults: A Population-Based Multi-Cohort Study"

A Population-Based Multi-Cohort Study (Supplementary Material)

Yeongmin Kim1, Taesic Lee2*, Chang-Myung Oh1*

1Department of Biomedical Science and Engineering, Gwangju Institute of Science and Technology, Gwangju, Korea

2Department of Family Medicine, Yonsei University Wonju College of Medicine, Wonju, Korea

*Correspondence should be addressed to:

Chang-Myung Oh, MD, PhD

Tae Sic Lee, MD, PhD

Supplementary Tables

Supplementary Ta b l e 1. General characteristics of the participants according to Fatty liver disease in KoGES (Korean Genome

Epidemiology Study)

Supplementary Table 2. Selection of multivariable for multiple logistic regression analysis

Supplementary Figures

Supplementary Figure 1. KNHANES cohort inclusion and exclusion diagram

Supplementary Figure 2. NHANES cohort inclusion and exclusion diagram

Supplementary Figure 3. KoGES cohort inclusion and exclusion diagram

Supplementary Figure 4. The percentage of Fatty Liver Status in KNHANES, KoGES , and. NHANES HSI 36, NHANES HSI 41.

Supplementary Figure 5. The prevalence of Diabetes, Dyslipidemia, and Hypertension in the decade in KNHANES and in the two decades in NHANES

Supplementary Figure 6. The relationship between chronic metabolic diseases and alcohol consumption in KNHANES (A) and NHANES cohort (B).
